# Custom Biomedical FAIR Data Analysis in the Cloud Using CAVATICA

**DOI:** 10.1101/2024.06.27.24309340

**Authors:** Seth R Berke, Kanika Kanchan, Mary L Marazita, Eric Tobin, Ingo Ruczinski

## Abstract

The historically fragmented biomedical data ecosystem has moved towards harmonization under the findable, accessible, interoperable, and reusable (FAIR) data principles, creating more opportunities for cloud-based research. This shift is especially opportune for scientists across diverse domains interested in implementing creative, nonstandard computational analytic pipelines on large and varied datasets. However, executing custom cloud analyses may present difficulties, particularly for investigators lacking advanced computational expertise. Here, we present an accessible, streamlined approach for the cloud compute platform CAVATICA that offers a solution. We outline how we developed a custom workflow in the cloud, for analyzing whole genome sequences of case-parent trios to detect sex-specific genetic effects on orofacial cleft risk, which required several programming languages and custom software packages. The approach involves just three components: Docker to containerize software environments, tool creation for each analysis step, and a visual workflow editor to weave the tools into a Common Workflow Language (CWL) pipeline. Our approach should be accessible to any investigator with basic computational skills, is readily extended to implement any scalable high-throughput biomedical data analysis in the cloud, and is applicable to other commonly used compute platforms such as BioData Catalyst. We believe our approach empowers versatile data reuse and promotes accelerated biomedical discovery in a time of substantial FAIR data.

## Introduction

Cloud computing has long held the promise of facilitating large-scale bioinformatic analyses due to its ability to house and assess big data ^1^. But as data expanded in the cloud, many available biomedical datasets were siloed and some were separated into distinct Data Coordinating Centers (DCCs), each with their own niche: GTEx for tissue-specific gene expression^2^, Gabriella Miller Kids First (GMKF)^3^ for childhood cancer and structural birth defects, exRNA for extracellular RNA^4^, and many more. These data silos presented an interoperability bottleneck for analyses in the cloud^5^. Academia, industry, and publishers recognized the incompatibility of much of the biomedical data landscape and together published the findable, accessible, interoperable, and reusable (FAIR) data principles which aimed to improve data access and sharing ^6^. While there is still room for improvement, FAIRness was largely a success as many data resources have embraced the principles and there continue to be increasing FAIRification efforts ^7^. One program especially facilitating this effort is the National Institute of Health’s (NIH) Common Fund Data Ecosystem (CFDE), an endeavor to harmonize multiple DCCs under a single umbrella ^8^. Although increased data ecosystem interoperability would seemingly imply expedited biomedical discovery, researchers have faced significant friction in the cloud, especially for those interested in carrying out specialized analyses.

Biomedical cloud data analysis platforms, also called data commons ^9^, are providers that aim to facilitate analysis by centralizing data storage and analytic tools to carry out workflows in the cloud. Many standard software packages (such as VCFtools ^10^ for analyses of sequencing data) are commonly made available by such providers, however due to the nonstandard nature of custom analyses, many tools (defined as distinct processing steps in an overall workflow) needed for their execution are often not pre-installed by the platforms ^11^. This has become especially true as providers continue to differentiate into niches with certain data and tools, leaving no singular platform versatile to every analysis. Thus, one platform that may include a needed specialized tool for a given pipeline may be incompatible with the rest of a researcher’s workflow or data ^11^, often leaving investigators reliant on local or high performance computing (HPC) clusters to carry out custom steps. The mixture of local, HPC, and cloud architecture to execute nonstandard workflows has led to an unwieldy technique where researchers scatter to adapt workflows (or portions of workflows) to architecture-specific complexities and gather results back together post-hoc. The issue pervades biomedical research; thus, the FAIR Principles for Research Software (FAIR4RS) have been established to improve the landscape of biomedical software analysis by making analytic tools more accessible and shareable ^12^. Therefore, there is an urgenct need for streamlined approaches to build comprehensive, customized workflows in the cloud that ensure the accessibility and implementation of specialized analytic tools.

We here describe such an approach, employing a CAVATICA workflow on FAIR data from the NIH Common Fund’s GMKF Pediatric Research Program (”Kids First”). On our local HPC we had previously downloaded WGS data from case-parent trios, ascertained on children with non-syndromic orofacial cleft (OFC) at birth, to assess and meta-analyze sex-specific effects on OFC risk using custom software available in the Bioconductor package trio^13^. The previously analyzed data sets were trios of participants with European, Central/South American, and Asian ancestry. Recently, a large number of case-parent trios of Filipino origin were sequenced through GMKF and uploaded onto CAVATICA. To avoid copying the data to our cluster we containerized our computational pipeline, ported it to CAVATICA, generated the workflow, executed the code, and returned the results for inclusion into the meta-analysis, which significantly increased the overall sample size. The approach is solely based on software containers, plus tool and workflow creation in the cloud. In this manuscript we detail the respective steps and offer guidance to enable investigators with basic computational skills to migrate, develop, and conduct their custom analyses in the cloud. Our approach is language and software package agnostic and is applicable to other commonly used compute platforms such as BioData Catalyst with access to the Database of Genotypes and Phenotypes (dbGaP) data sets. The Summary Statistics of dbGaP Data webpage shows that the cumulative counts of approved data access requests is approaching 100,000 in the year 2024, making our approach very timely to avoid thousands of local copies of the data being generated. We discuss the CAVATICA infrastructure that enables the development and housing of biomedical research studies and emphasize the platform’s adherence to common software development practices such as version control, documentation, and debugging for code review ^14^. In addition, we offer suggestions on how to minimize time and cost during workflow development and execution. We illustrate our workflow approach through our case study, investigating sex-specific genetic effects on OFC risk, and show how the CAVATICA analysis on the Filipino trios contributed to identifying a genetic marker exhibiting sex-specific effects at a genome-wide significance level.

## Results

A CAVATICA research study centers around a Project Space that provides an intuitive entry point to house a particular investigation. For our analysis we first crafted a custom workflow to clean the genomic data, and then another workflow to carry out a genome-wide association study to identify sex-specific loci underlying OFC risk. Here, we describe our case study in more detail, and the necessary steps for other investigators to create their own our custom analytic workflows to assess FAIR data in the cloud.

### The Case Study

OFCs are the most common congenital craniofacial anomalies and the second most common birth defect worldwide. OFCs can be categorized into non-syndromic malformations, which occur in isolation, and syndromic malformations, which occur with another malformation or as part of a recognized malformation syndrome. Isolated or non-syndromic OFCs (including cleft lip, cleft palate, and cleft lip with cleft palate) vary between the sexes and birth prevalence rates differ across populations. OFCs are commonly categorized into two anatomically and embryologically distinct entities based on embryologic and epidemiologic patterns: cleft lip with or without cleft palate (CL/P) and cleft palate alone (CP) ^15^. In particular, non-syndromic CL/P, the phenotype we analyze here, occurs more frequently in males than females (male to female ratio approximately 2:1) ^16^. To elucidate the underlying mechanisms for differential risk to CL/P between the sexes, we analyzed whole genome sequences from case-parent trios generated through the GMKF Program. We performed genotypic transmission disequilibrium tests (gTDTs) using our in-house Bioconductor package trio^13^ to test for interactions between autosomal SNPs and sex.

The GMKF trios were sequenced at the Broad Institute and Variant Call Format (VCF) files were disseminated by the GMKF Data Resource Center (DRC). In the past, dissemination usually meant investigators obtaining a copy of the data using a file transfer protocol and storing the data locally. We had previously downloaded and analyzed WGS data from trios of European (289 trios), Central/South American (259 trios) and Asian (124 trios) ancestry on our local computing cluster. To avoid the practice of making data copies, we decided to analyze a more recently released set of trios of Filipino ancestry (370 trios) directly in its cloud environment on CAVATICA. The first step of every such analysis is to run quality control (QC) procedures on the VCF files to obtain the analyzable data. We used VCFtools ^10^ to remove multi-allelic sites and indels, filter sites with labels other than ‘PASS’, and to remove variants and individual genotypes with low mean depth and low quality scores (see Methods for details). We then created binary PLINK^17^ files (.bed, .bim and .fam) from these cleaned VCFs to integrate the pedigree information, and removed variants with low minor allele frequency (MAF) and those significantly deviating from Hardy Weinberg Equilibrium (HWE), statistics calculated from the parental genotypes. Trios with one or more samples found during QC to exceed a missingness threshold or producing a large number of Mendelian errors and were removed from the analysis. Relatedness of subjects was verified using identity-by-descent estimates for all pairs of individuals calculated from linkage disequilibrium (LD) pruned common variants. In all, we identified 16 trios to be dropped from the original 370 based on quality control.

A total of 354 Filipino ancestry trios with 7,866,804 variants passing all QC criteria were analyzed on CAVATICA using the gTDTs with a SNP main effect and a SNP*×* sex interaction term. The full analysis including functional annotation of variants will be published elsewhere (manuscript in preparation), we here simply show one example how the CAVATICA cloud analysis on the Filipino trios contributed to identifying a genetic marker exhibiting sex-specific effects at a genome-wide significance level. In brief: the 1df test for the SNP *×* sex interaction was nominally significant for SNP rs2715199 (located on chr2 at position 87,896,956, build hg38, A/G alleles) in all populations with the same direction of effect (e.g., the parameter for the interaction in the gTDT conditional logistic regression model). In the European trios the effect was estimated to be 1.59 (p = 4.6 *×*10*^−^*^3^, MAF = 5%), in the Central/South American trios the effect was estimated to be 1.32 (p = 9.9 *×*10*^−^*^3^, MAF = 7%), and in the Asian trios the effect was estimated to be 2.80 (p = 4.3 *×*10*^−^*^2^, MAF = 3%). The strongest association however was observed in the Filipino trios, where the effect was estimated to be 3.24 (p = 5.2 *×*10*^−^*^6^, MAF = 5%). In the resulting meta-analysis of the four populations using inverse variance weighting of effects, implemented in the program METAL^18^, the interaction parameter estimate was 1.78 with a p-value of 5.7 *×*10*^−^*^8^. After genomic-control correction^19^, this SNP achieved a genome-wide significant difference between the sexes of p = 1.1 *×*10*^−^*^8^.

The QC related tools on CAVATICA (Figure 1) are described below with technical detail. The actual command line code and the scripts used in the tools are available from the manuscript GitHub page.

- Basic QC Filtering | VCFTools Removing multi-allelic sites and indels, filtering sites with labels other than ‘PASS’, and removing variants and individual genotypes with low mean depth and low quality scores. The input is the multi-sample VCF file released by the sequencing center containing all complete trios. The cleaned output file, labeled in the grey circle as Basic QC-ed vcf is used in the subsequent calculation of other QC statistics and also as input for the gTDT analysis pipeline.
- PLINK File Creation | PLINK Creating binary PLINK files (.bed, .bim and .fam) to integrate the pedigree information into the QC steps for this family-based association study.
- Summary Stats | PLINK Calculating variant statistics such as missing genotype calls per variant across all samples, and variant MAF and HWE (calculated from the parents only). Variants that exhibit high genotype missingness, low MAF, or are significantly out of HWE are not included in the association analysis. Also calculating percent missing genotype calls for each sample. The output .lmiss and .imiss files contain the information to determine which samples are dropped due to their high genotype missingness.
- Variant Filtering | PLINK Generating updated PLINK files after removing trios with at least one sample showing high genotype missingness, and filtered on variant missingness, MAF and HWE. The input file Drop High Missingness Samples.txt was derived from the .lmiss and .imiss files.
- Mendelian Error | PLINK Calculating the percentage of Mendelian errors in the trios (proband genotypes that are incompatible with parental genotypes under Mendelian inheritance, assuming no *de novo* events) and generating a list of trios to be dropped from the gTDT analysis due to high error rates.
- LD Pruning Part 1| PLINK Deriving LD blocks of SNPs from the PLINK files, to be used in the following pruning step.
- LD Pruning Part 2 | PLINK Creating pruned binary PLINK files from LD block information for subsequent quality control.
- PLINK File Merge | PLINK Merging together pruned single-chromosome PLINK files into a whole-genome PLINK file.
- Heterozygosity | PLINK Calculating sample SNP heterozygosity rates to detect potential issues such as sample contamination.
- IBS | PLINK Calculating identity-by-descent estimates between pairs of samples to verify relatedness as given in the pedigree files.

**Figure 1:**
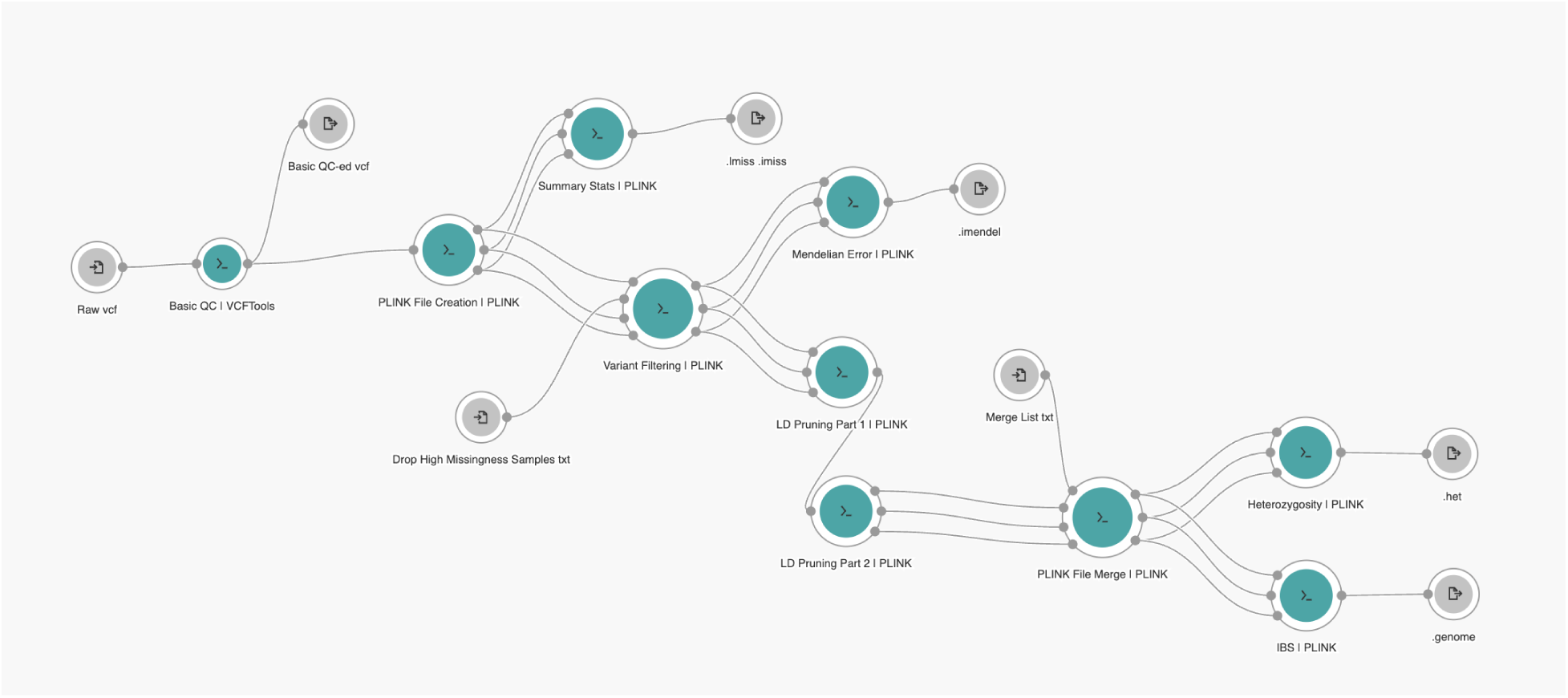
A CWL schematic of the QC pipeline in the workflow visual editor. Teal colored circles represent CAVATICA tools. Grey circles represent input and output files. Lines indicate the connection between files and tools.

The gTDT related analysis tools on CAVATICA (Figure 2), described in more detail:

- Variant Level QC | VCFTools Reading the VCFs generated after basic QC, removing the trios failing QC, updating variant statistics (missingness, MAF and HWE) after removing these trios, and removing variants based on these updated statistics.
- gTDT | R + Bioconductor Executing the gTDT with a SNP *×* sex interaction as implemented in the trio Bioconductor package. For this sex-specific case-parent trio analysis, the tool also reads the sex and pedigree information available in the metadata.
- MAF Percentage | VCFTools Extracting the MAF information for the output file.
- MAF Column Creation | R + SplitStackShape Selecting either the REF or ALT allele as the minor allele based on observed allele frequencies, and creating the MAF column for the output file.
- Column Extraction | BCFTools Extracting information using BCFTools ^20^ for the final results output file, including chromosome, genomic position, rs number, REF and ALT alleles.
- Merge | R + Bioconductor Merging all pieces of information into the final results output file.

**Figure 2:**
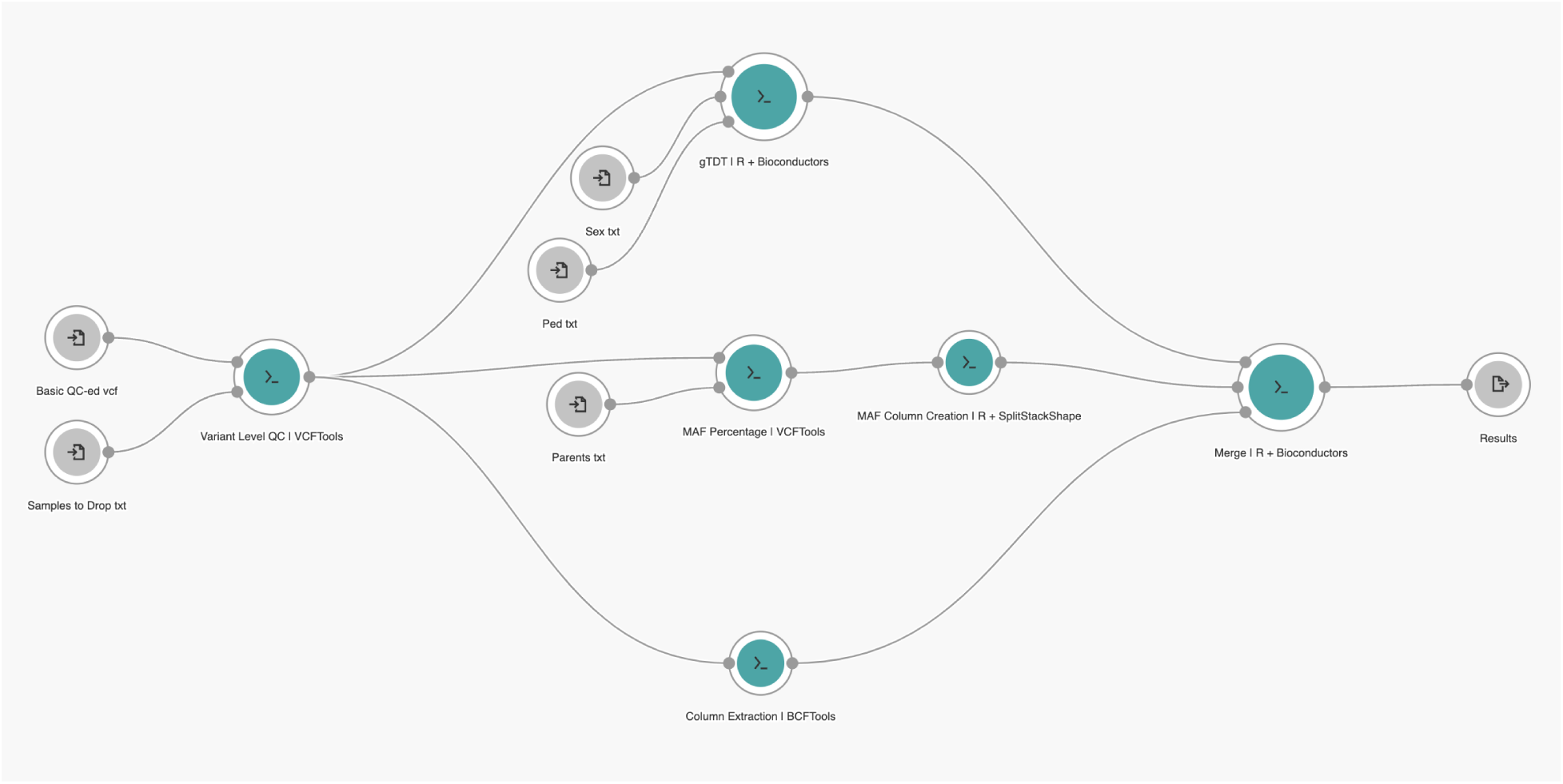
A CWL schematic of the gTDT analysis pipeline in the workflow visual editor. Teal colored circles represent CAVATICA tools. Grey circles represent input and output files. Lines indicate the connection between files and tools.

### Creating Custom Workflows

We here present the fundamental concepts (Figure 3) needed to carry out a specialized cloud computing analysis with big data in CAVATICA, such as the one described above. We also offer narrated videos on a companion website that help to demonstrate how to implement various steps. The videos are not hosted on the manuscript’s GitHub page due to file size limitations, instead a link to the companion website has been added.

**Figure 3:**
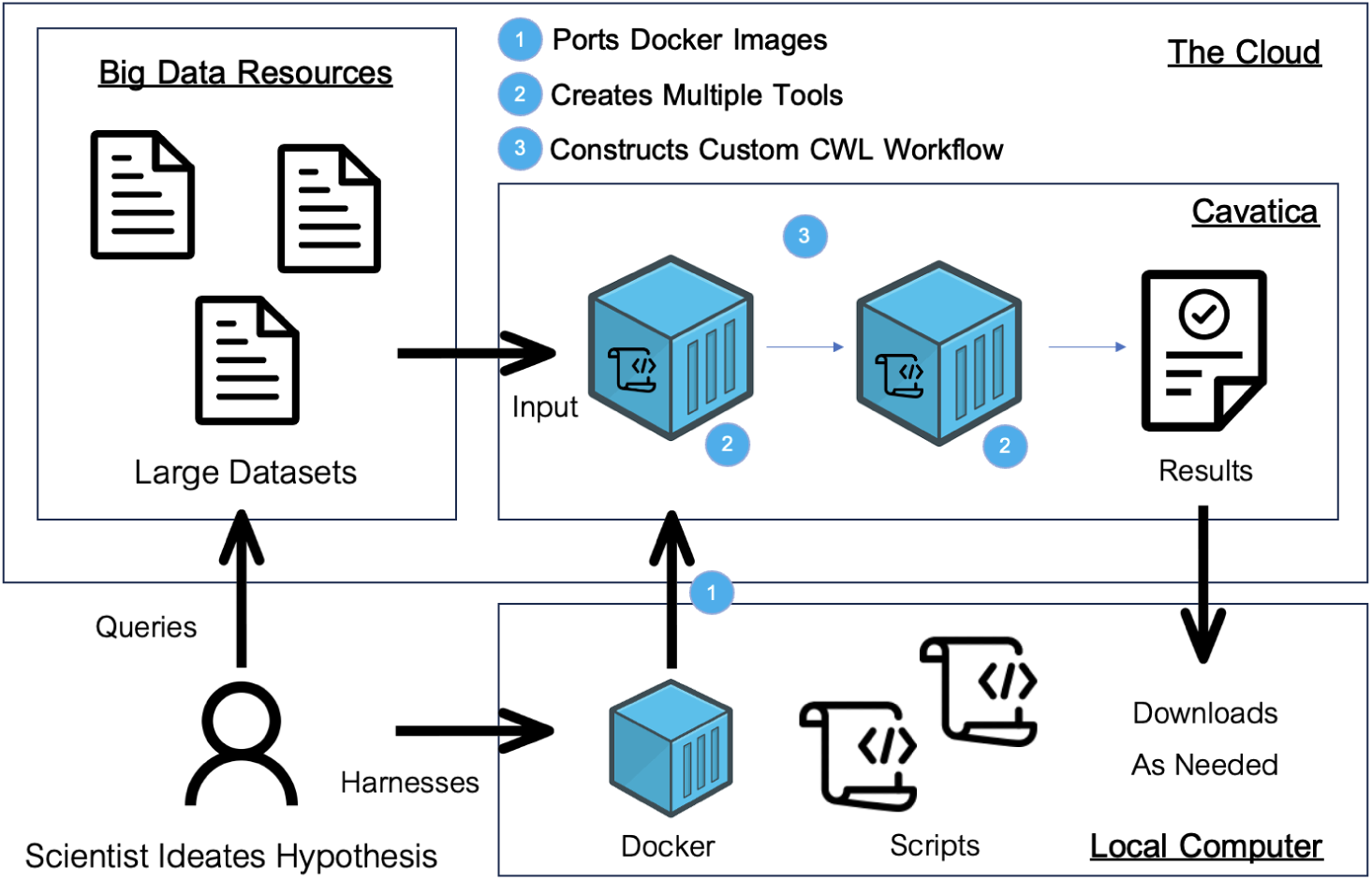
A schematic of the process to carry out custom cloud workflows. Details of the process and data access considerations are described in detail in the Methods section.

#### Docker

Software containers comprise the necessary software and dependencies for a given computational job into light-weight, portable environments. They have gained traction over virtual machines which require more computational resources^21^. Docker arguably has become the industry standard for software containerization. It streamlines container development with three key components: the i) Dockerfile, which holds the instructions to construct a ii) Docker Image, which is a snapshot of the software and dependencies that will be run in the iii) Docker Container, which is the associated executable environment. It is important to note that Docker Images are immutable, ensuring enduring reproducibility in computational research^22^. Here is the typical structure of a custom Dockerfile:

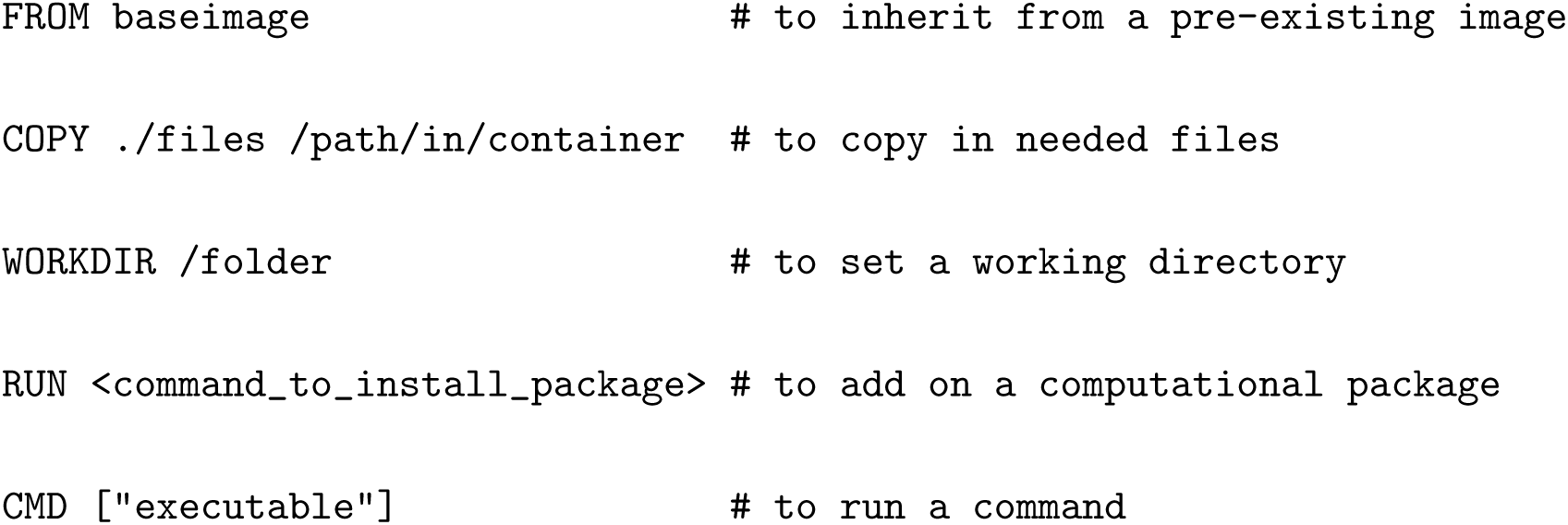

Docker is open source, allowing published Docker Images from the software development community to be freely pulled from the DockerHub. Docker is also modular, which means that images with only parts of the required software can be added to fulfill all software requirements. In our application, we used several Docker Hub Images built on top of others to port our polylingual, multi-software pipeline onto CAVATICA. Specifically, we used Docker Hub Images for VCFTools and BCFTools. However, there was no pre-existing Docker Container with the Bioconductor package trio for the gTDT analysis, which we had previously carried out locally in the statistical environment R. Thus, we crafted our own Dockerfile on top of the Bioconductor base image, as shown below. Naturally, this method is extendable to any

Bioconductor package.

~~~
FROM bioconductor/bioconductor_docker:devel-amd64
RUN R -e “install.packages(‘data.table’)
RUN R -e ‘BiocManager::install(“VariantAnnotation”, force = TRUE)’
RUN R -e ‘BiocManager::install(“trio”)’
~~~

The Dockerfile is then transformed into a portable Docker Image and pushed to CAVATICA’s Docker Registry. The companion website contains a video how to create such a Docker Image and push it to CAVATICA. In the below, “name” is equal to the Docker registry repository a user must create in CAVATICA.

~~~
docker login pgc-images.sbgenomics.com
docker build -t name . --platform=“linux/amd64”
docker tag name pgc-images.sbgenomics.com/username/name
docker push pgc-images.sbgenomics.com/username/name
~~~

The complex computational requirements necessitated by custom workflows can be resolved through Docker. In CAVATICA, each tool is execucted within a Docker Container, a notable difference from the popular prior methodology of a comprehensive Docker Container to containerize an entire pipeline. By tackling containerization on a more granular level, CAVATICA allows for more control over tool and workflow creation.

#### Tools

A CAVATICA tool is a fundamental building block of a complex analysis workflow, serving as a standalone executable for a distinct processing step in the overall pipeline. Each tool is executed within a specified Docker Container that includes its required software. A tool is depicted graphically in CAVATICA’s workflow Visual Editor as a teal circle with input and output ports (Figures 1 and 2). Researchers can choose from publicly available pre-installed tools, or they may choose to create their own. The companion website to this manuscript contains videos that reinforce the descriptions here on how to create a custom CAVATICA tool and how to create a CAVATICA tool from pre-installed software.

To customize tools for specific analyses, investigators first start by naming the tool which then gives them access to the editor for creation. The editor has a fill-in interface to craft commands and specifications. Researchers must select a Docker Image that contains the proper software dependencies, construct an executable bash command, include file scripts when needed, and configure inputs and output specifications. There also exists an option to save output and error logs generated during runtime for debugging. The tool editor implements software development standard practices that any integrated development environment (IDE) would, such as version control similar to Git, debugging aid and error logs, and extensive documentation opportunities. Here, we highlight as an example the tool that ran our custom gTDT analysis, termed gTDT | R + Bioconductor (Figure 2). The script gTDT.R, which is available on our GitHub page, was added as a supplementary executable file to the tool. Each input and output file in the bash command corresponds to a grey circle in the visual editor. For example, Ped txt is an input for the gTDT | R + Bioconductor tool (teal circle). Once constructed, the tool’s command line code reads:

~~~
Rscript ./gTDT.R input_ped.ext input_sex.ext input_vcf.ext > stdout.out
~~~

Importantly, standalone tool development presents the opportunity to combat the often costly tweaking of bioinformatic pipelines (”tinkering”) in the cloud. To reduce cost while testing our workflow adaptation to the cloud, we leveraged a few computational principles: i) verifying the efficacy of each tool as a standalone step, ii) conducting testing with minimally sized data, and iii) leveraging memoization, an optimization technique for the re-running of computational jobs that re-uses results for the aspects of a task that remain unedited. Through this approach, we streamlined tool validation with minimal pricing. For our full gTDT analysis (Figure 2) we created six distinct tools as described above, each of which has their Dockerfile and source code published on our GitHub. We tested each tool with our three “tinkering” steps and confirmed the efficacy of each. While standalone execution helps with development, running an entire analysis one step at a time creates inefficiencies: intermediate, derived files clutter storage and the execution of many tasks takes significant manual labor and time. Therefore, crafting together these modular steps into a polylingual workflow becomes necessary for efficient research.

#### CWL Workflow System

Workflow Systems are implemented to weave together distinct tools into an overall analytic pipeline. In CAVATICA, the standard system is the Common Workflow Language (CWL), a workflow language with comprehensive polylingual workflow support, portability, and compliance with regulatory requirements ^23^. While CWL is often cited as having low ease of use as a workflow manager ^24^, CAVATICA eases this concern through a visual drag-and-drop editor such that workflows can be crafted without the explicit use of the tedious syntax of the underlying CWL code. Users add custom made or publicly available tools to the editor and connect tools through their input and output ports, creating lines between tools that represent the flow of files within the pipeline (Figures 1 and 2). The workflows editor also maintains software development standards such as version control, documentation, and extensive debugging. Leveraging memoization is an especially important step here, as researchers can avoid rerunning an entire complex pipeline by reusing the results of unchanged workflow tools. The companion website has a video how to create such a CAVATICA workflow from the individual tools.

While carrying out a workflow task users must select initial input files, and when there are multiple files for the same input that would necessitate multiple runs, a “Batch Task” may be created. In doing so, multiple runs of the workflow are initialized and will be parallelized over each file. For example, the 22 single autosome multi-sample VCF files were processed simultaneously. CAVATICA automatically schedules jobs without the need for investigators to write bash logic to optimize workflow execution. Users can select the “Stats and Logs” panel of their workflow execution to inspect information on tools in runtime alongside their time and computational resource allocation. In the “Instance Metrics” panel, users can assess closely the Central Processing Unit (CPU) and Random Access Memory (RAM) load their tasks demand. By observing where the computational resources of their chosen instance type (a virtual server for cloud computing) exceed the demands of a given task, users can optimize time and cost by selecting a more suitable instance. Amazon Web Services (AWS) offers a variety of instances with different CPU/RAM ratios that can be selected for a given tool, enabling researchers to achieve more granular optimization. In all, the visual editor and options available allow for efficient workflow development and execution.

## Discussion

In this manuscript we presented a generic approach for specialized CAVATICA cloud analyses and illustrated the approach by analyzing whole genome sequences of case-parent trios to detect sex-specific genetic effects underlying OFC risk. We note that our approach is also applicable to other platforms such as BioData Catalyst, and we hope that the guidance offered in this manuscript will enable other investigators with basic computational skills to implement and execute their own analytic pipelines in the cloud. In particular, we believe our approach adheres to the FAIR data principles by promoting data reuse for new biomedical discovery.

The development process - once understood - was quick in its implementation and efficient in its execution. We used standard software development approaches for code development and porting. Version control was especially critical in saving progress for each modular step; we found that the ability to return to a specific time point during development was both useful and an ease of mind for the case of post-modification workflow breakdown. We implemented the discussed methodology to “tinker” without excessive time or cost by periodically testing each single-step tool with minimal data sizes. This allowed us to re-run tools during the debugging process. Leveraging the CWL workflow system and the automatic job scheduler allowed us to carry out complex workflows with ease, keeping the costs low for a highly scalable analytic pipeline.

Developing this approach admittedly involved a somewhat steep learning curve and overcoming some hurdles. Developing cloud computing with a polylingual workflow logic requires a solid knowledge of the fundamentals for software containment, tool creation, and workflow language systems. A new user might become frustrated when encountering a lack of documentation about the computing infrastructure and the file system. In particular with our videos on the companion website we hope we will be able to flatten that learning curve for other investigators. Once understood, we believe the implementation of any study can become very much streamlined. Our goal was to lower the entry point for cloud research by empowering researchers from diverse scientific backgrounds to craft cloud-based custom pipelines and to investigate novel hypotheses with varied datasets.

Another hindrance we faced was the lack of support files for our gTDT analysis, for example pedigree and phenotype files with the sex of the probands. As is unfortunately quite common in the biomedical data ecosystem, we had to contact the principal investigator of the study to obtain these metadata. We strongly believe that DCC integration should include a more comprehensive file support system with all study information readily available and accessible, in accord with the FAIR data principles, to allow for diverse biomedical analyses. The well known issue of metadata non-standardization within the field is arguably among the largest obstacles for true FAIR data usage.

Metadata standardization will also be critical to harness the full potential the access to multiple large data resources offers. The approach presented in this manuscript is extendable to other SevenBridges technologies such as the Cancer Genome Cloud containing the The Cancer Genome Atlas and BioData-Catalyst with access to dbGaP data sets. The analytic techniques are interoperable, and any CAVATICA pipeline can be executed in CGC or BDC, and vice versa. We hope that the guidance we offer will be useful for investigators who wish to develop or migrate specialized analyses for specific use cases on the cloud.

In all, with the abundance of FAIR data and increased interoperability between diverse data sets and types from different DCCs, we are enthusiastic about the potential for novel scientific hypotheses to be raised and tested within many large datasets now cloud-accessible. We believe there could be a real paradigm shift especially for “non-standard” analyses, which are typically carried out in-house on local machines or HPC computing.

## Methods

### The Genotypic Transmission Disequilibrium Test with an Interaction

The allelic transmission disequilibrium test (aTDT) proposed by Spielman^25^ is commonly used in the analysis of case–parent trio data to test for an association of a genotype and a disease. The aTDT quantifies whether an allele at a genetic locus was preferentially transmitted from the parents to the affected offspring. In contrast, the genotypic transmission disequilibrium test (gTDT) considers for each family all four possible pairs of parental alleles at a locus and compares the genotypes observed for the affected probands to the pseudo-controls, i.e. the unobserved genotypes of the other Mendelian children.

Conditional logistic regression is then applied to these strata, each consisting of the genotypes of the affected proband and the three matched pseudo-controls^26^. The gTDT has several advantages over the aTDT. It allows to select a specific genetic mode of inheritance and can be more powerful than the aTDT^27^. The gTDT further generates parameter estimates and standard errors (in addition to just p-values in the aTDT) which allows for more straightforward meta-analyses, in particular when results from family and population based studies are to be combined ^28^. Lastly, the gTDT allows testing for interactions between SNPs and other covariates such as sex to assess effect modifications.

The main drawback of the gTDT compared to the aTDT is its potentially much higher computational burden since iterated weighted least squares are commonly employed to obtain the parameter estimates in the conditional likelihood ^29^, which can be prohibitive when tens of millions of variants have to be assessed. We previously showed however that Mendel’s laws however impose a structure on the genotypes in the gTDT that allows us to derive exact closed-form solutions for the parameter estimates of the conditional logistic regression models when testing for an additive, a dominant, or a recessive effect of a SNP, and we also showed that such analytic parameter estimates exist when considering interactions between SNPs and binary covariates such as sex ^30^. Our procedure, implemented in the freely available Bioconductor package trio, is based on a simple counting / tabulation procedure and avoids iterative numeric optimization, reducing computing time by two orders of magnitude. This software package allowed us to analyze tens of millions of markers for their association with oral clefting in our meta-analysis, and to assess whether genetic effects differ between the sexes.

For the QC procedures, we used VCFtools to remove multi-allelic sites and indels, filter sites with labels other than ‘PASS’, remove variants with mean depth values less than 10X or quality value less than 20, and remove genotype calls with less than 10X depth or quality less than 20. We then created binary files (.bed, .bim and .fam) from these cleaned VCFs to integrate the pedigree information of the affected probands using PLINK. We excluded the variants with more than 20% missingness, minor allele frequency below 1%, and those significantly deviating from Hardy Weinberg Equilibrium with a p-value less than 10*^−^*^6^. Trios were excluded from the analysis if they had at least one sample with more than 2% missing genotypes, showed ambiguous relatedness based on pairwise IBD estimates, or had an excessive number of Mendelian errors.

### Project Spaces on CAVATICA

A CAVATICA project space integrates five key components of biomedical data research which are commonly handled with disparate tools in local environment or a high performance computing cluster. The components are referred to on CAVATICA as “Dashboard”, “Files”, “Apps”, “Tasks” and “Data Studio” (Figure 4).

**Figure 4:**
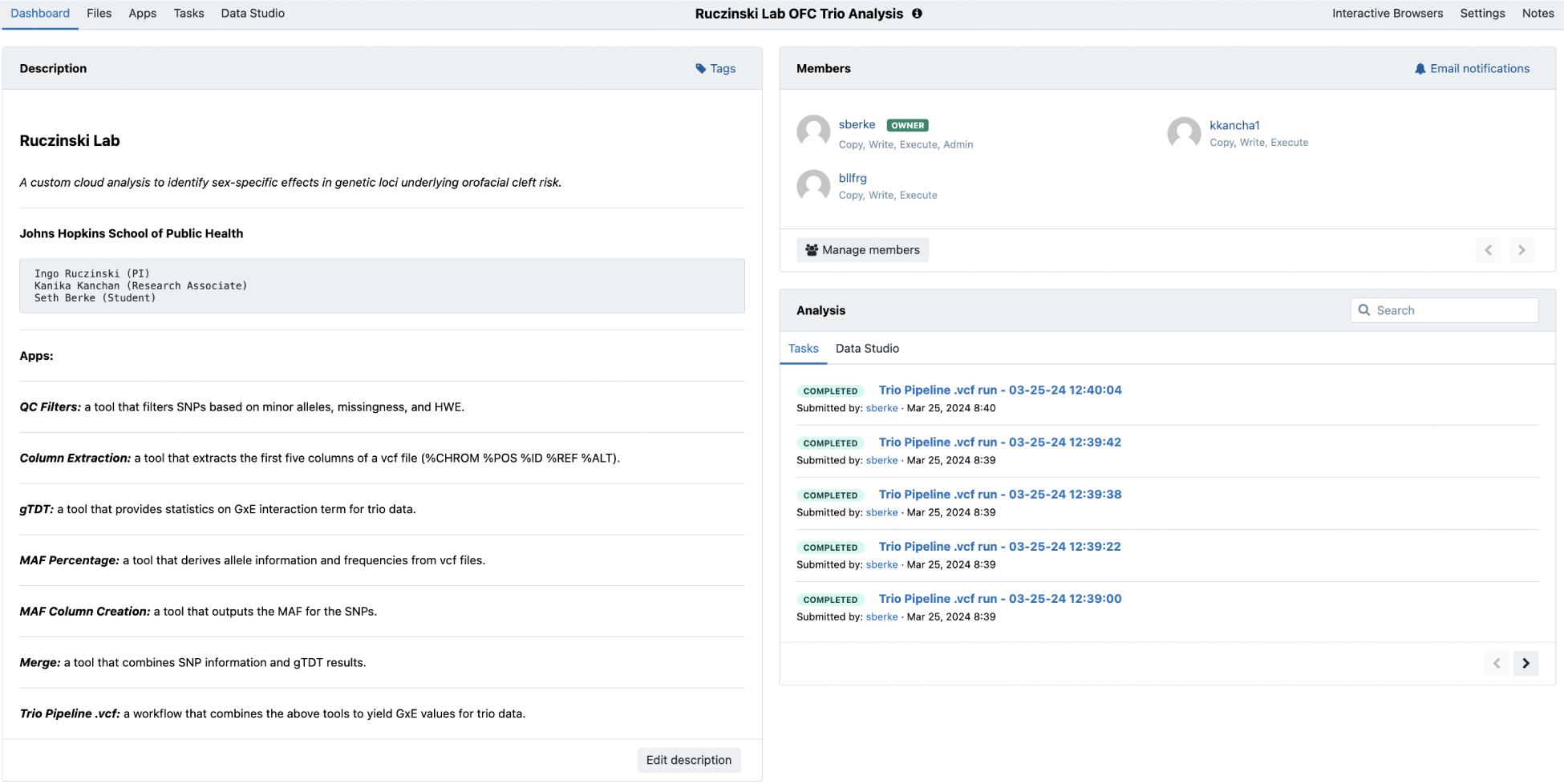
A CAVATICA Project Space is the portal to a research project. Here, files are stored, member permissions are managed, tools and workflows are created, and computational tasks are executed.

#### Dashboard

The Dashboard represents a comprehensive view of a team’s research. It includes a Markdown “Description” panel to describe the project aims. With the “Manage Users” subsection, the project administrator can control the user privileges (copy, write, execute). This extends the simplicity of user-access management techniques from other more commonly known cloud-based collaborative software, such as Google Documents. A final section shows the status of task executions.

#### Files

The Files section allows for a variety of methods for data uploading, and users can choose to upload data from diverse sources. Importing is facilitated from the Global Alliance for Genomics and Health (GA4GH) Data Repository Service (DRS) Application Program Interface (API). Through this technique, file manifests can be derived for example from DCC data and uploaded. The process is light-weight, as it leverages each file’s Uniform Resource Identifier (URI) hyperlink, which allows for file storage through an access link to the original source without having to copy the data. URIs are both unique and immutable, which help them identify the data location and access protocol ^31^. Researchers may also choose to import datasets from local computers via direct upload and from HPC Clusters via the SevenBridges’ File Transfer Protocols (FTPs). The data ecosystem on CAVATICA maintains FAIR principles: F) every file has a persistent identifier (PID) and the capacity to include extensive metadata specifications, A) these files are retrievable by their PID within any project space on the platform, I) data is easily integrated from diverse sources and types for novel investigations, and R) files are optimized for reuse and data URIs allow for original data to be reused numerous times. More details on data access are given in the separate section “Data Access on CAVATICA” below.

#### Apps

A CAVATICA app falls under one of two categories: a tool or workflow. Within the “Apps” section, the user may choose to import an already existing app, create a new app, or modify an already built app. A tool is a distinct processing step while a workflow is a combination of multiple tools woven together. The app creation protocol maintains software development standards: version control, documentation, and debugging. Through app creation, users can craft polylingual CWL workflows customized for any analysis pipeline.

#### Tasks

Tasks in CAVATICA are computational jobs that execute either a tool or a workflow. Each is accompanied by information about its status (”Draft”, “Queued”, “Running”, “Aborted”, “Failed”, “Completed”), its job initiator, its execution date, its duration, and its price. Task management is maintained by CAVATICA infrastructure which acts as a Portable Batch System (PBS) that eliminates the need for bash scripting for execution. While drafting a task, a user can choose to use spot instances to reduce the cost of the task, implement memoization to re-use pre-computed tasks where applicable, and run batch tasks to compute multiple files in parallel. During the execution of a task, researchers can investigate the “Stats and Logs” page to see the time and resource allocation of a tool or overall workflow. In this space, researchers also may see error logs to aid in the debugging of their code. Finally, there is an “Instance Metrics” panel to assess computation load on the CPU, RAM, input / output loading (IO), and more.

#### Data Studio

The Data Studio houses JupyterLab and RStudio as computational environments within which users can initiate sessions to directly interact with their data, derived files, or their results. During this session, researchers may develop analytic scripts in Python or R, respectively, that later can be crafted into a tool or implemented in an overall workflow. The data and visualizations produced from these scripts within the native Data Studio environment may also be downloaded into the Project Space.

### Data Access on CAVATICA

Access to data is critical for the success of any research project. Here we describe the mechanisms in place to ensure secure and compliant data handling on CAVATICA. The platform integrates with multiple data repositories and follows stringent protocols to manage user access. The data access mechanisms on CAVATICA support a wide range of genomic research activities and uphold the required standards of data security and compliance.

CAVATICA employs the Researcher Auth Service (RAS) to streamline access to controlled datasets. RAS is a federated authentication service that verifies researchers’ identities and their right to access specific datasets, especially those containing human genetic information. This system ensures that all data access on CAVATICA complies with the ethical standards and privacy regulations required by research governance. RAS on CAVATICA is coordinated via the NIH eRA Commons system, a system for tracking roles of researchers and governing data access permissions. Through integration with the dbGaP, CAVATICA also allows researchers to access these genomic and phenotypic data. Access to dbGaP is controlled and requires users to have appropriate authorization, which is facilitated through the RAS. This ensures that sensitive data is only accessed by qualified individuals.

CAVATICA facilitates interoperability to enhance the utility of data across various platforms and tools. By adhering to common standards and protocols including those set by GA4GH, CAVATICA ensures that data imported from or exported to other platforms can be integrated without compatibility issues. This approach supports a collaborative and efficient research environment where data can be shared and utilized across different systems and studies. One of the core components of this interoperability is CAVATICA’s DRS, a GA4GH standard. The DRS provides a uniform API for accessing data objects across cloud environments, regardless of the data’s location. This means that researchers can retrieve data from GMKF and other compliant platforms using a consistent set of tools and protocols. This is particularly beneficial for projects that span multiple data types and sources, as it reduces the complexity and overhead associated with data management. Furthermore, using a DRS enabled file provides a symbolic link to that data’s original storage location in the cloud, and checks access requests for every call against user permissions. This means data already stored elsewhere in the cloud are not billed against the user’s project. When data are accessed, the platform performs analysis in the same local cluster where the data are stored, further enhancing security.

The DRS server also facilitates the movement of data within the platform and ensures that data uploaded to projects on CAVATICA are readily available on other platforms such as the Cancer Genomics Cloud (CGC) and BiodataCatalyst (BDC). In addition, CAVATICA also hosts a range of datasets that are accessible through its Data Explorer tool, including datasets from GMKF and The Cancer Genome Atlas (TCGA). This feature allows researchers to search, filter, and access datasets based on various parameters such as disease and data type. The Data Explorer is a tool for researchers to quickly find the necessary data for their research projects, streamlining the process of data discovery and access.

## Data availability

The Filipino WGS trio data are available in the *Genomics of Orofacial Clefts in the Philippines* workspace on CAVATCA and on dbGaP under study accession number phs002595.v1.p1. Approval for this study was given by the following Ethics Committees: University of the Philippines, Manila, Research Ethics Board (FWA00018728); the University of Iowa Institutional Review Board (FWA00043007, IRB approval ID# 200812719); the University of Pittsburgh Institutional Review Board (FWA00006790, IRB approval ID# STUDY19080127). Informed consent was obtained for all participants using a certified translation into Tagalog of the English consent form approved by the study coordinating center at the University of Pittsburgh.

## Code availability

All relevant code is freely available from the first author’s manuscript GitHub page.

## Data Availability

The trio data are available in the Genomics of Orofacial Clefts in the Philippines workspace on CAVATCA and on dbGaP under study accession number phs002595.v1.p1

https://www.ncbi.nlm.nih.gov/projects/gap/cgi-bin/study.cgi?study_id=phs002595.v1.p1

## Acknowledgments

S.B., K.K., M.L.M and I.R. were supported by NIDCR R01-DE031855. This work was also supported in part by the Division of Intramural Research, National Institute of Allergy and Infectious Diseases, NIH. Data collection was supported by R01-DE016148, and sequencing by X01-HD100701. The authors thank Sarah W Curtis and Elizabeth Leslie for supplying necessary metadata files and information for the cloud data analysis.

## Author Contributions

S.B. and I.R. conceived the workflow. K.K. developed code used on a local cluster that was also ported to CAVATICA as part of the analysis. E.T. provided guidance on navigating the CAVATICA environment. S.B. implemented the CAVATICA workflow, conducted the analyses, and created the videos on the manuscript companion website. M.L.M advised on the genetic data analysis. All authors reviewed the manuscript.

